# Early recanalization among patients undergoing bridging therapy with tenecteplase or alteplase

**DOI:** 10.1101/2023.05.08.23289701

**Authors:** Thomas Checkouri, Gaspard Gerschenfeld, Pierre Seners, Marion Yger, Wagih Ben Hassen, Nicolas Chausson, Stéphane Olindo, Jildaz Caroff, Gaultier Marnat, Frédéric Clarençon, Jean-Claude Baron, Guillaume Turc, Sonia Alamowitch, the TETRIS and PREDICT-RECANAL collaborators

**Author notes:** **Corresponding author:** Prof. Sonia Alamowitch. Service des Urgences Cérébro-Vasculaires. Hôpital Pitié-Salpêtrière, 47-83 boulevard de l’Hôpital, 75651 Paris Cedex 13, France. Drs Checkouri and Gerschenfeld contributed equally.

## Abstract

**Background:** Intravenous thrombolysis (IVT) with alteplase or tenecteplase prior to mechanical thrombectomy (MT) is the recommended treatment for large-vessel occlusion acute ischemic stroke (LVOS). There are divergent data on whether these agents differ in terms of early recanalization (ER) rates before MT, and little data on their potential differences in terms of established ER predictors such as time elapsed between IVT and ER evaluation (IVT-to-ER_eval_ time), occlusion site and thrombus length.

**Methods:** We compared the likelihood of ER after IVT with tenecteplase or alteplase in anterior circulation LVOS patients from the PREDICT-RECANAL (alteplase) and TETRIS (tenecteplase) French multicenter registries. ER was defined as a modified thrombolysis in cerebral infarction score 2b-3 on first angiographic run or non–invasive vascular imaging (magnetic resonance or computed tomography angiography) in patients with early neurological improvement. Analyses were based on propensity score overlap weighting (PSOW, leading to an exact balance in baseline characteristics between the treatment groups) and confirmed with adjusted logistic regression (sensitivity analysis).

**Results:** A total of 1865 patients were included. ER occurred in 156/787 (19.8%) and 199/1078 (18.5%) patients treated with tenecteplase or alteplase, respectively (odds ratio, 1.09 [95%CI 0.83–1.44]; *P*=0.52). A differential effect of tenecteplase *vs* alteplase on the probability of ER according to thrombus length was observed (*P*_interaction_=0.003), with tenecteplase being associated with higher odds of ER in thrombi > 10 mm (odds ratio, 2.43 [95% CI 1.02-5.81]; *P*=0.04). There was no differential effect of tenecteplase *vs* alteplase on the likelihood of ER according to the IVT-to-ER_eval_ time (*P*_interaction_=0.40) or occlusion site (*P*_interaction_=0.80).

**Conclusion:** Both thrombolytics achieved ER in a fifth of LVOS patients, with potentially greater effect of tenecteplase in larger thrombi. There was no significant differential influence of IVT-to-ER_eval_ time or occlusion site on likelihood of ER.

## INTRODUCTION

In patients with large-vessel occlusion acute ischemic stroke (LVOS), early and complete recanalization is associated with improved functional outcome.^1^ Bridging therapy, which consists in intravenous thrombolysis (IVT) followed by mechanical thrombectomy (MT), is the recommended treatment to achieve timely recanalization in this population.^2,3^ The role of IVT in LVOS management has been recently questioned by six randomized controlled trials aiming to demonstrate the non-inferiority of a bypass strategy omitting IVT before MT.^4–9^ While two trials found MT alone to be non-inferior to bridging therapy,^8,9^ the others did not show its non-inferiority.^4–7^ A recent study-level meta-analysis of these trials failed to demonstrate non-inferiority based on margins of 1.3% or even 5%,^10^ but results from individual patient data meta-analysis are still pending. One potential advantage of bridging therapy is IVT-induced early recanalization (ER) before MT, which may be associated with better functional outcome.^11^ Several groups have explored factors associated with ER after IVT with alteplase,^12,13^ identifying three major predictors: (i) more distal occlusion site; (ii) longer time elapsed between IVT and evaluation of ER (IVT-to-ER_eval_ time); and (iii) thrombus properties such as thrombus length or perviousness.^12–14^

Tenecteplase is a genetically modified tissue–plasminogen activator which has improved fibrin specificity and longer half-life, allowing single-bolus administration, whereas alteplase dose is administered along a 60-minute infusion.^15,16^ Tenecteplase has recently been recommended over alteplase before MT for LVOS in European guidelines,^2,3,17^ following the demonstration of higher ER rates and better functional outcomes in the Tenecteplase versus Alteplase before Endovascular Therapy for Ischemic Stroke (EXTEND-IA TNK) trial.^18^ Subsequently, tenecteplase use in the LVOS population has increased and several “real–world” studies have reported reassuring efficacy and safety data, with functional outcome and intracerebral hemorrhage rates similar to those of alteplase.^19–23^ Reduced process times have also been reported, owing to its simpler administration.^24–26^ More recently, two non-randomized studies reported higher ER rates with tenecteplase compared to alteplase,^27,28^ while in the Alteplase compared to Tenecteplase (AcT) trial, both thrombolytics achieved similar ER rates.^29^ Hence, there are conflicting data on whether tenecteplase actually yields higher ER rates than alteplase, and little data on potential differences in terms of ER predictors between the two agents. Further knowledge on these points is needed, as any potential difference in efficacy between them could have significant implications in clinical practice.

In this study, we aimed to compare the likelihood of ER before MT among two French multicentric registries of LVOS patients treated with alteplase or tenecteplase, and the potential influence of the established predictors of ER, namely IVT-to-ER_eval_ time, occlusion site and thrombus length.^12^

## METHODS

The data that support the findings of this study are available from the corresponding author upon agreeing to a data sharing agreement. This article follows the STROBE reporting guidelines.

### Study Population

We used two French multicenter databases of patients intended for bridging therapy after demonstration of LVOS by magnetic resonance imaging (MRI) or computed tomography angiography (CTA). In both groups, patients were referred for MT either at a comprehensive stroke center (CSC) following direct admission, or after secondary transfer if they received IVT at a primary stroke center (PSC). Data were collected from stroke centers between May 2015 and March 2017 (alteplase 0.9 mg/kg group: PREDICT–RECANAL cohort, 8 CSC and their 23 referring PSC)^12^ and from May 2015 to June 2021 (tenecteplase 0.25 mg/kg group: TETRIS cohort, 3 CSC and 2 PSC).^21^

Patients who fulfilled the following criteria were included: (i) 18 years or older, (ii) evidence of LVOS of the anterior circulation, defined as an occlusion of either the intracranial internal carotid artery (ICA), the first (M1) or second (M2) segment of the middle cerebral artery (MCA), (iii) IVT within 4.5 hours of symptoms onset, or in the presence of a magnetic resonance imaging (MRI) mismatch between diffusion-weighted imaging (DWI) and fluid-attenuated inversion recovery (FLAIR) in case of unknown onset and (iv) evaluation of ER within 180 minutes from IVT (see below).

This study was approved by the Sorbonne University Research Ethics Committee (CER– 2021–1053). As per current French law regarding retrospective studies of anonymized standard care data, patients were informed of their participation in this research and offered the possibility to withdraw. No written consent was required for this research.

### Clinical Data

The following variables were extracted from both registries: age, sex, vascular risk factors (high blood pressure, diabetes mellitus, smoking, history of myocardial infarction and stroke), pre-stroke medication (antiplatelets, anticoagulants, statin), neurological severity measured with the National Institutes of Health Stroke Scale (NIHSS) score on admission, time between symptom onset and start of IVT (onset-to-IVT time) and IVT-to-ER_eval_ time.

### Imaging Data

Patients underwent brain MRI on admission (or CTA in case of contraindication to MRI), as recommended by current French guidelines.^30^ Acute stroke MRI protocols slightly varied across centers but included DWI, FLAIR, intracranial time-of-flight angiography and a T2* or susceptibility-weighted sequence. In PREDICT-RECANAL, an experienced stroke neurologist reviewed the pre-IVT imaging of all included patients.^12^ In TETRIS, all pre-IVT imaging of included patients were reviewed by experienced stroke neurologists or neuroradiologists.^21^ In both cohorts, pre-IVT imaging reviewers had at least 5 years of experience in stroke treatment. The following variables were collected: (i) intracranial occlusion site: ICA, M1 dichotomized as proximal or distal based on the MCA origin-to-clot interface distance (<10 and ≥10 mm, respectively), and M2 defined as starting after the main MCA bifurcation; (ii) length of the susceptibility vessel sign (SVS), measured on T2*-MRI or susceptibility-weighted imaging, as previously published; (iii) DWI lesion extent using the Alberta Stroke Program Early CT Score (DWI–ASPECTS); and (iv) presence of a tandem cervical ICA occlusion. We chose not to measure thrombus length and infarct size on CT given the limited number of patients who underwent CT in both cohorts, and the lack of appropriate way to pool these CT-based variables with corresponding MR-based variables in the same statistical analysis.

### ER evaluation

The primary outcome was substantial ER defined as a modified Thrombolysis in Cerebral infarction (mTICI) score ≥ 2b on the first DSA run for the intended MT. Complete ER (mTICI 3) was the secondary outcome. In patients with early neurological improvement, mTICI score was evaluated on the non–invasive vascular imaging (MR angiography or CTA) performed to reevaluate the need for MT in the PSC or CSC. Pre-interventional mTICI was evaluated in comparison with the pre-IVT site of occlusion, as in EXTEND–IA TNK.^18^ ER evaluators had access to both imaging to this end. In both cohorts, brain DSA, CTA or MRA were evaluated for ER by experienced (at least five years) stroke neurologists or neuroradiologists, who differed from the one who interpreted the imaging initially.

### Statistical analysis

Quantitative variables were described as mean ± standard deviation (SD) or median (interquartile range [IQR]), as appropriate, and qualitative variables as counts and percentages. For our main analysis, in order to account for imbalance in potential confounders for the association between thrombolytic treatment and ER, we chose to use a propensity-score overlap weighting (PSOW) approach,^31^ after multiple imputations (n=20) for missing data, performed under the missing-at-random assumption.

Because a sizeable proportion of patients had no visible SVS,^32^ which precluded to account for their thrombus length, we derived two PSOW models, one based on the whole cohort and the other on the subgroup of patients who had a visible SVS. All baseline variables (except for MRI-specific variables for the first PSOW model) were included in the logistic models used to estimate the propensity score of each patient to be intended for tenecteplase (as opposed to alteplase), considering potential multicollinearity. Balance of baseline characteristics between the two treatment groups was assessed before and after PSOW by calculation of absolute standardized mean differences (ASMD). An ASMD ≤ 10% was interpreted as a negligible difference.^33^ It is noteworthy that for all included baseline variables, PSOW leads to an exact balance (i.e., ASMD=0%) between the treatment groups without excluding any patients.^31^

The association between treatment group and ER was estimated through odds ratios (ORs) and their 95% confidence intervals (95% CIs), calculated in univariable conditional logistic regressions. The ORs from each imputed dataset were combined using Rubin’s rules. Potential heterogeneity in treatment effect depending on predefined variables (occlusion site, IVT-to-ER-evaluation time, and thrombus length)^12^ was assessed in conditional logistic models with calculation of *P* values for interaction (*P*_interaction_). Before conducting these analyses, we non-parametrically examined the possibly nonlinear relationship between variables of interest (IVT-to-ER_eval_ time; thrombus length) and ER with restricted cubic splines. IVT-to-ER_eval_ time and thrombus length were also analyzed as categorical variables (0-29 min, 30-59 min, 60-119 min, ≥120 min and <10 mm *vs* ≥10 mm, respectively).

To assess the robustness of our results, we conducted sensitivity analyses based on adjusted logistic regression, without propensity score or imputations for missing data. A bidirectional stepwise method was used for the selection of variables to be included in the logistic model. The selected variables were age, NIHSS score on admission, time period (before or after 2016), onset-to-IVT time, occlusion site, type of transfer (secondary transfer to a CSC *vs* direct admission), diabetes mellitus, and type of imaging for ER evaluation (DSA *vs* other).

All tests were two-sided and the significance level was set at *p*<0.05. Analyses were performed using SAS 9.4 (SAS Inc). GT and TC had full access to all the data in the study and take responsibility for its integrity and the data analysis.

## RESULTS

### Whole cohort analysis

Over the study period, 1865 patients were included, 787 in the tenecteplase and 1078 in the alteplase group (**Figure 1**). **Table 1** summarizes patients’ baseline characteristics in the two treatment groups. Before PSOW, several meaningful differences (ASMD > 10%) were observed: compared with the alteplase group, patients in the tenecteplase group were older, more frequently using anticoagulants, less frequently received IVT at a PSC and were imaged with MRI, had a longer onset-to-IVT and a shorter IVT-to-ER_eval_ time, and more frequently underwent DSA.

**Table 1.**
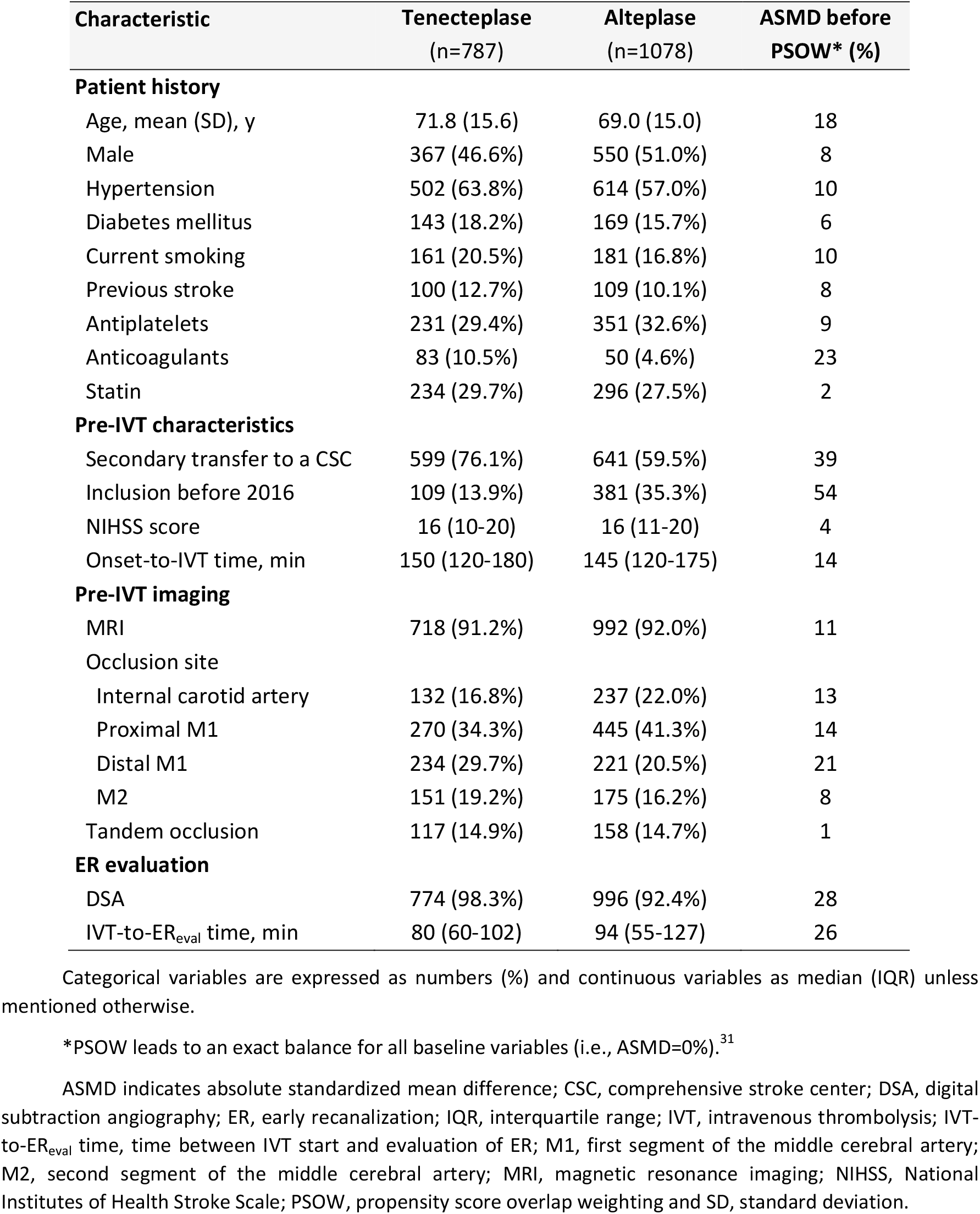
Baseline Characteristics (whole cohort, PSOW)

**Figure 1.**
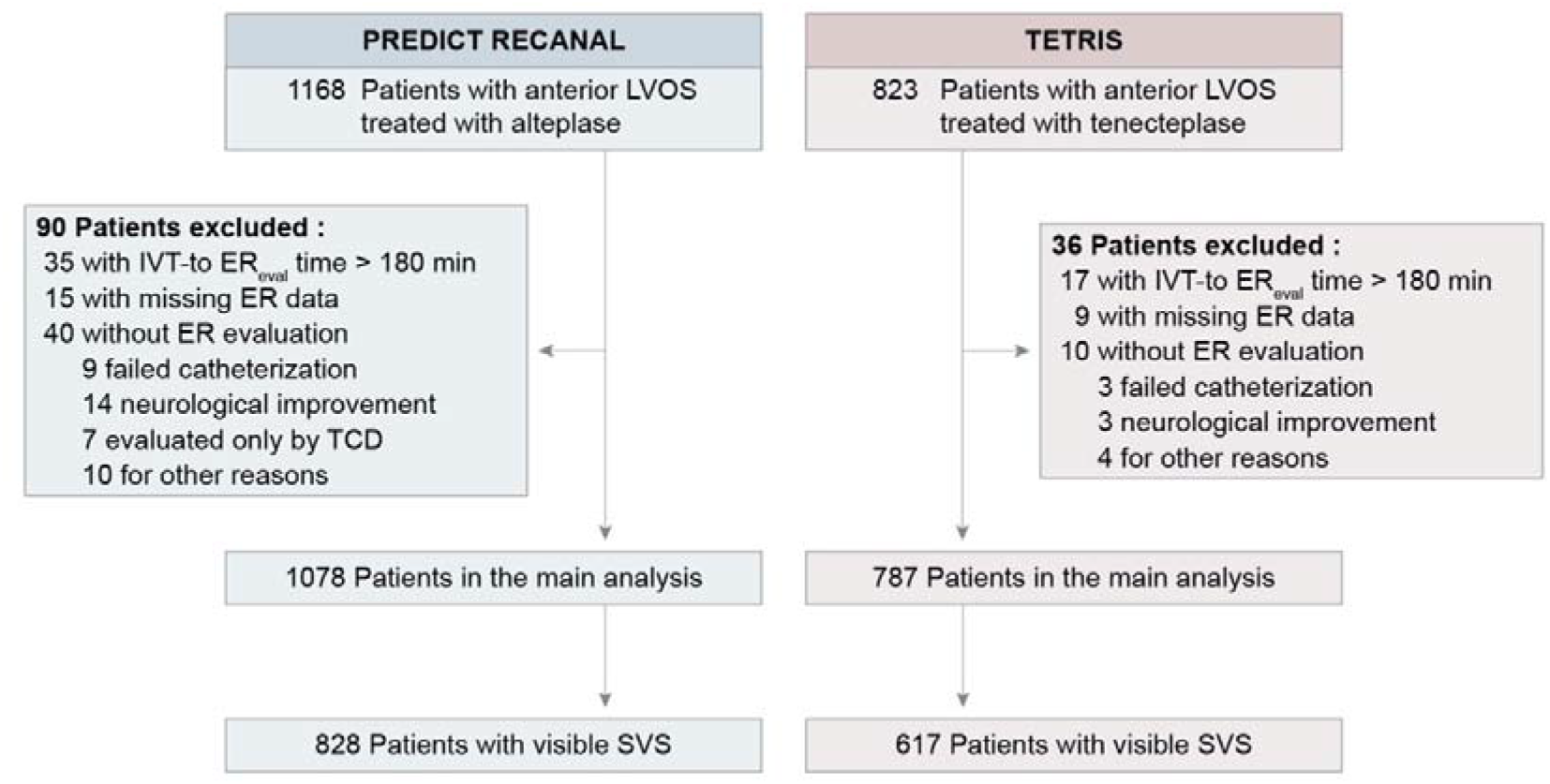
Flow Chart. ER indicates early recanalization; IVT-to-ER_eval_ time, time between IVT start and evaluation of ER; LVOS, large-vessel occlusion acute ischemic stroke; SVS, susceptibility vessel sign.

For the main analysis, a first PSOW model was derived after multiple imputation for missing values, in which all patients were included. Both treatment groups were exactly balanced (ASMD = 0%, see Methods) with regards to all baseline variables. ER occurred in 156 (19.8%) and 199 (18.5%) patients treated with tenecteplase or alteplase, respectively (OR 1.09 [95% CI 0.83-1.44]; *P*=0.52). There was no influence of the type of imaging used to assess ER (DSA *vs* MRA or CTA; *P*_interaction_=0.95). Complete ER (mTICI 3) occurred in 26 (3.3%) and 52 (4.8%) patients treated with tenecteplase or alteplase, respectively (OR 1.16 [95% CI 0.67-1.99]; *P*=0.59). There was no differential effect of treatment on the probability of ER across IVT-to-ER_eval_ times, either considered as a continuous (*P*_interaction_=0.40) or a categorical variable (*P*_interaction_=0.28; **Figure 2**). ER rates were similar with tenecteplase or alteplase according to occlusion site (*P*_interaction_=0.80**; Figure 3**).

**Figure 2.**
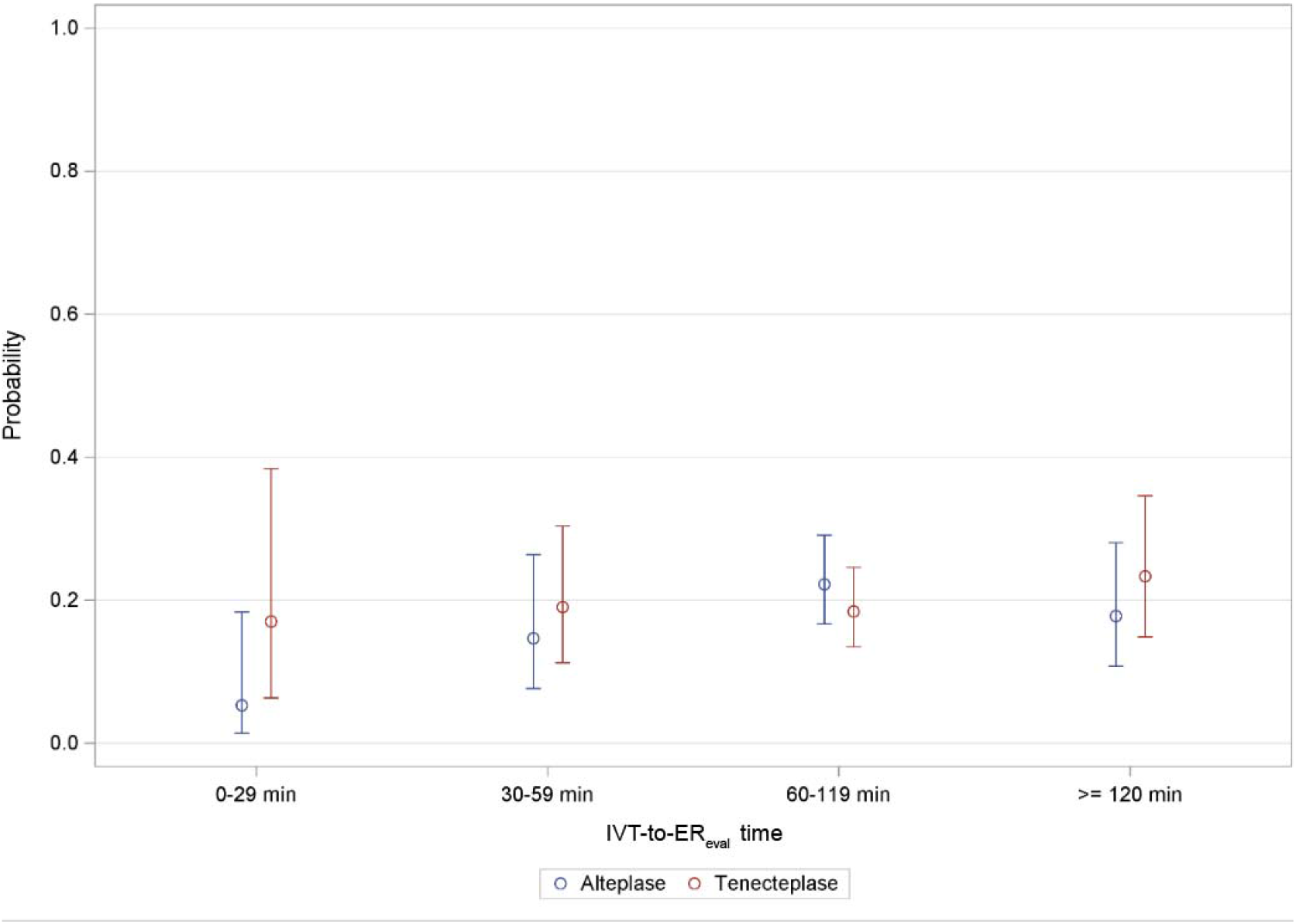
Probability of early recanalization in patients treated with tenecteplase or alteplase, according to IVT-to-ER_eval_ time. ER indicates early recanalization; IVT-to-ER_eval_ time, time between IVT start and evaluation of ER.

**Figure 3.**
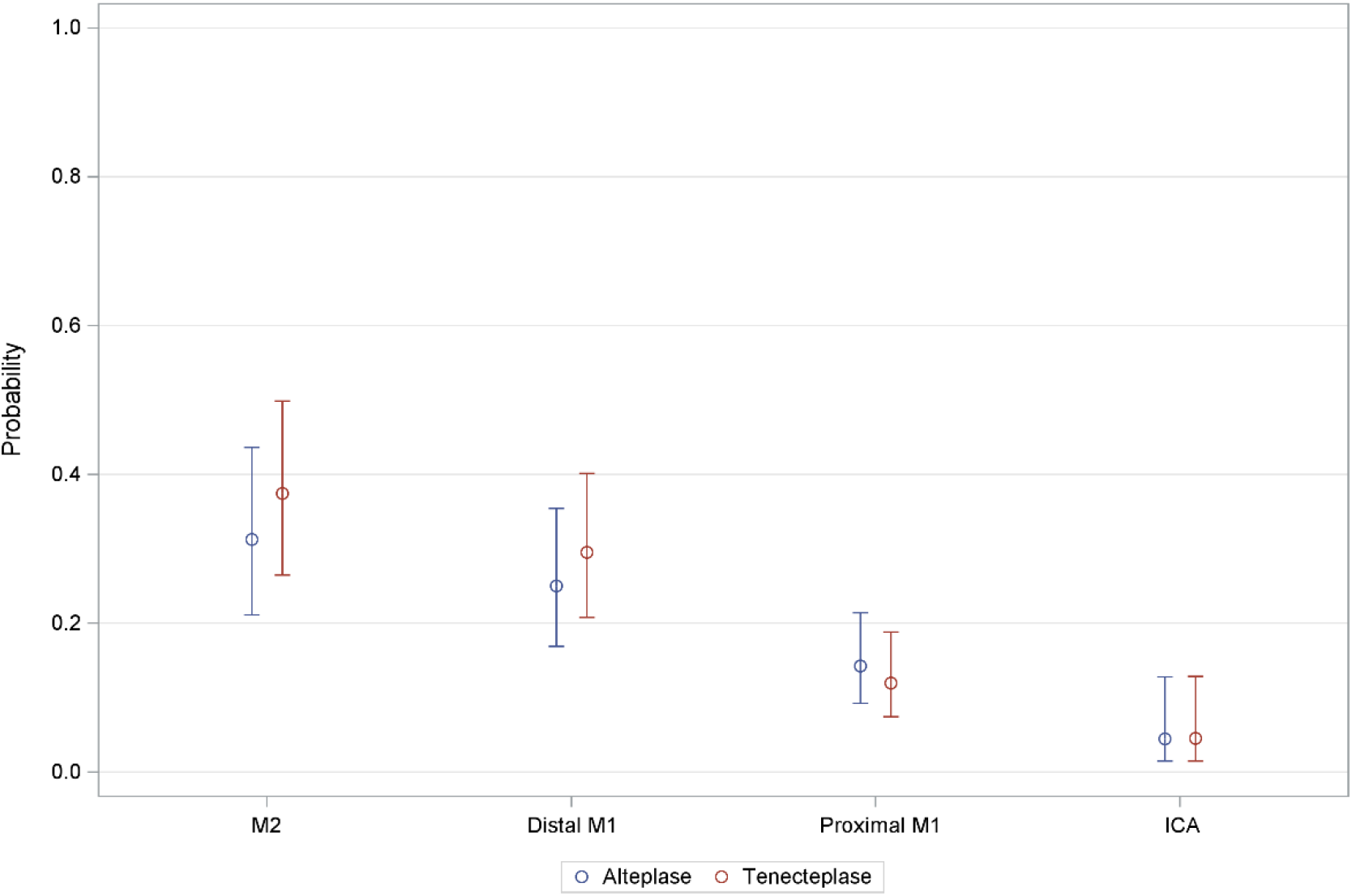
Probability of early recanalization in patients treated with tenecteplase or alteplase, according to the occlusion site. ICA indicates internal carotid artery; M2, second segment of the middle cerebral artery; M1, first segment of the middle cerebral artery.

We observed similar results in sensitivity analyses based on adjusted logistic regression. Variables included in the multivariable model were age, NIHSS score on admission, inclusion period (before 2016), onset-to-IVT time, occlusion site, secondary transfer to a CSC, diabetes mellitus, and ER evaluation imaging modality. The probability of ER was not significantly different in patients treated with tenecteplase *vs* alteplase (adjusted OR 1.10 [95% CI 0.82-1.46]; *P*=0.54). Complete ER did not also differ in patients treated with tenecteplase *vs* alteplase (adjusted OR 1.17 [0.63-2.14], *P*=0.62). There was no differential treatment effect on the probability of ER across IVT-to-ER_eval_ times (*P*_interaction_=0.67), nor by occlusion site (*P*_interaction_=0.79).

### Subgroup of patients with visible SVS

We derived a second PSOW model for the subgroup of patients who had visible SVS on pre-treatment MRI (n=1445), which led to an exact balance in baseline characteristics between the two groups (**Table S1**). We observed a differential effect of tenecteplase *vs* alteplase on the probability of ER according to thrombus length considered as a continuous variable (*P*_interaction_=0.003; **Figure 4**). This was also true (*P*_interaction_=0.001) if SVS length was dichotomized (SVS < 10 mm: OR 0.74 [95% CI 0.41-1.33], *P*=0.31; SVS ≥ 10 mm: OR 2.43 [95%CI 1.02-5.81], *P*=0.04).

**Figure 4.**
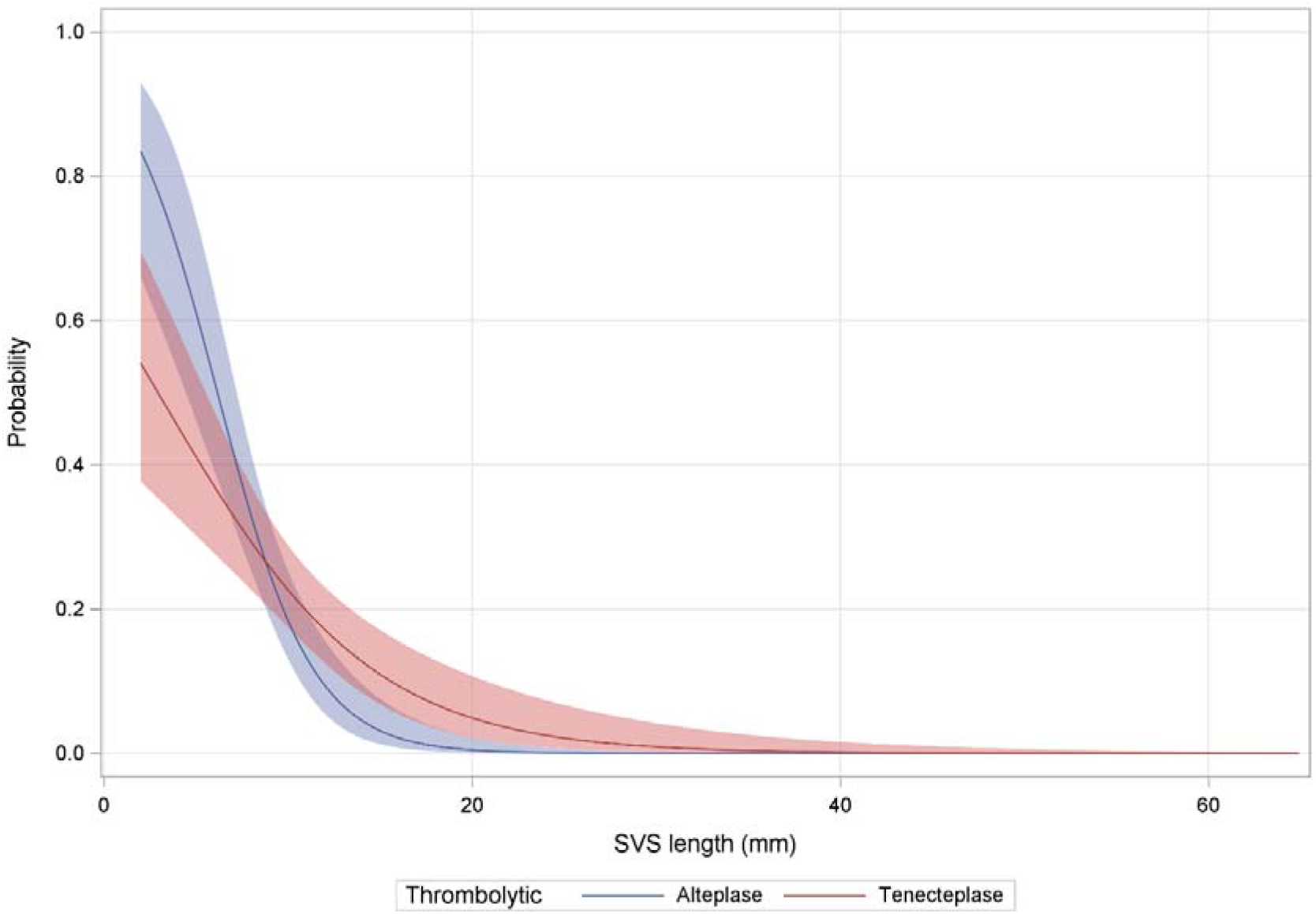
Probability of early recanalization in patients treated with tenecteplase or alteplase, according to thrombus length. SVS indicates susceptibility vessel sign.

Similar results were observed in sensitivity analyses based on adjusted logistic regression, with a differential treatment effect according to SVS length (*P*_interaction_<0.0001).

## DISCUSSION

In this study, we found no significant difference in odds of ER between tenecteplase and alteplase in the whole cohort. However, tenecteplase use was associated with a higher likelihood of ER for larger thrombi (≥ 10 mm). Occlusion site and IVT-to-ER_eval_ time affected ER rates similarly for alteplase and tenecteplase.

The benefit of IVT before MT remains currently debated, following diverging results from six IVT bypass non-inferiority trials, which almost exclusively used alteplase and were conducted in CSCs. ER is among the main postulated mechanisms to explain a potential benefit of IVT, as supported by two key observations: (i) shorter onset-to-reperfusion times are associated with better functional outcome;^34^ and (ii) even when incomplete, ER is associated with better functional outcomes.^35,36^ However, available data from clinical trials and registry studies comparing ER rates between tenecteplase and alteplase are divergent. Thus, while higher ER rates were achieved with tenecteplase than alteplase in the EXTEND-IA TNK trial (22% and 10%, respectively, median IVT-to-ER_eval_: 55 minutes), this was not the case in the AcT trial (around 10% for both thrombolytics, median IVT-to-ER_eval_: 38 minutes).^18,29^ Notably, reported ER rates with alteplase in IVT bypass studies were even lower (1 to 7%), with median IVT-to-ER_eval_ time ranging from 24 to 34 minutes.^4,5,7–9^

Our analysis found similar ER rates among patients treated with tenecteplase and alteplase (19.8% and 18.5%, respectively), with a median IVT-to-ER_eval_ time of 88 minutes. This finding is in line with reported ER rates with alteplase in patients with IVT-to-ER_eval_ times >80 minutes,^12,37,38^ and with tenecteplase in the pooled analysis of the EXTEND-IA TNK part 1 and 2 trials.^37^ We observed no differential effect of tenecteplase *vs* alteplase on the incidence of ER across IVT-to-ER_eval_ intervals in our analyses. This finding stands apart from some previous data that support that, while the ER rate quickly reaches about 20% with tenecteplase, even for short IVT-to-ER_eval_ times,^18,21,27,37,39^ it increases more slowly with alteplase.^28,39^ While Figure 2 seems visually in favor of tenecteplase among patients with short IVT-to-ER_eval_ times, the relevance of this trend is questionable given that non-linear association was checked for in the continuous variable and that the interaction analysis was negative. Further studies focusing on short treatment times will be required to explore the possible superiority or tenecteplase in this setting. The lack of statistical significance in our analysis could be driven by the higher proportion of patients secondarily transferred to a CSC and hence with longer IVT-to-ER_eval_ times in our cohort (around two-thirds). Indeed, secondarily transferred patients were fewer in both the EXTEND-IA TNK trial (about one-fourth) and a recently published pooled analysis of individual patient data from clinical trials and a prospective registry (about one-half).^18,28^ On this note, the results from the DIRECT-TNK IVT bypass trial with tenecteplase (NCT05199194) are awaited with high interest.

Regarding thrombus location and length, we found a strong association between likelihood of ER and occlusion site, as previously reported for both agents.^13,21,28,40,41^ Regarding SVS length, in a recent analysis of LVOS patients treated with alteplase, this variable was retained over occlusion site to predict lack of post–IVT recanalization.^12^ Our analysis demonstrates a favorable differential effect of tenecteplase in larger (≥ 10mm) thrombi. This result contrasts with a recent study which reported, using the clot burden score, higher odds of ER with tenecteplase only in patients with low clot burden.^41^ Clot burden score is a pragmatic approach to quantify thrombus load, and has been shown to be a good predictor for ER and 3-month clinical outcome.^42,43^ While it may indirectly reflect thrombus length, a recent study only found a weak correlation between SVS thrombus length and clot burden score.^43^ Hence, in the absence of additional data it is difficult to draw conclusions on this discrepancy. However, our finding would appear coherent with recently published results based on an *in vitro* human blood clot model, which found that tenecteplase was more effective against larger clots than alteplase.^44^ Further studies on this topic are needed, as a better efficacy of tenecteplase compared to alteplase in larger clots would be of particular importance as larger thrombi have been associated with early neurological deterioration and poor clinical outcomes.^45,46^

Our study has several strengths. It is based on two large multicentric LVOS patient registries with similar practices. Both databases had less than 1% missing data for each variable. We used a modern statistical approach, PSOW, which allows an optimal control of measured confounders without exclusion of patients. Our results remained robust when applying a second approach based on adjusted logistic regression.

Our study also has several limitations. First, it was based on retrospectively collected data, with unmeasured confounding factors that may influence our results. Second, we lacked additional data such as number of passes during MT, post–MT recanalization and functional outcome. Third, each cohort included patients treated over different time periods, and for instance most patients included in TETRIS were included after PREDICT-RECANAL had ended.^12^ This difference may have influenced some key metrics, as MT procedures became more widely implemented in routine clinical practice over time.^47^ Fourth, thrombus measurement was done manually, which may affect accuracy for smaller thrombi, and was centralized in PREDICT-RECANAL but not in TETRIS.

## CONCLUSION

Both tenecteplase and alteplase provide timely substantial ER in a fifth of LVOS patients who undergo bridging therapy. Our result favoring tenecteplase over alteplase among patients with larger thrombi may have implications for LVOS management. Other potential advantages of tenecteplase in bridging therapy remain to be studied.

## Data Availability

The data that support the findings of this study are available from the corresponding author upon reasonable request.

## Nonstandard Abbreviations and Acronyms

AcT: Alteplase compared to Tenecteplase trial
ASMD: absolute standardized mean difference
ASPECTS: Alberta Stroke Program Early CT Score
CTA: Computed tomography angiography
CSC: comprehensive stroke center
DSA: digital subtraction angiography
DWI: diffusion-weighted imaging
ER: early recanalization
EXTEND-IA TNK: Tenecteplase versus Alteplase before Endovascular Therapy for Ischemic Stroke trial
FLAIR: fluid-attenuated inversion recovery
ICA: internal carotid artery
IVT: intravenous thrombolysis
IVT-to-ER_eval_ time: time between intravenous thrombolysis start and evaluation of early recanalization
LVOS: large-vessel occlusion acute ischemic stroke
MCA: middle cerebral artery
MT: mechanical thrombectomy
NIHSS: National Institutes of Health Stroke Scale
mTICI: modified Thrombolysis in Cerebral infarction
PSC: primary stroke center
PSOW: propensity score overlap weighting
SVS: susceptibility vessel sign
TETRIS: Tenecteplase Treatment in Ischemic Stroke

## ONLINE ONLY SUPPLEMENTARY MATERIAL

Table S1. Baseline Characteristics of the subgroup of patients with visible SVS.

## ACKNOWLEDGMENTS

None

## APPENDIX

PREDICT-RECANAL collaborators: Caroline Arquizan, Yves Berthezene, Grégoire Boulouis, Serge Bracard, Nicolas Bricout, Tae-Hee Cho, Arturo Consoli, Vincent Costalat, Jean-Philippe Cottier, Séverine Debiais, Benjamin Gory, Hilde Henon, Bertrand Lapergue, Michael Obadia, Catherine Oppenheim, Fernando Pico, Michel Piotin, Sébastien Richard, Mathieu Zuber.

TETRIS collaborators: Clémence Blanc, David Calvet, Christian Denier, Sam Ghazanfari, Erwah Kalsoum, Yann L’Hermitte, Laurence Legrand, Jean-Sebastien Liegey, Malgorzata Milnerowicz, Olivier Naggara, Mathilde Poli, Kévin Premat, Igor Sibon, Candice Sabben, Eimad Shotar, Didier Smadja, Laurent Spelle.

